# Feasibility, Acceptability, and Barriers to Implementing Select Non-Pharmaceutical Interventions to Reduce the Transmission of Pandemic Influenza - United States, 2019

**DOI:** 10.1101/2021.09.22.21263963

**Authors:** Faruque Ahmed, Noreen Qualls, Shelly Kowalczyk, Suzanne Randolph Cunningham, Nicole Zviedrite, Amra Uzicanin

## Abstract

**Objective:** To assess feasibility and acceptability of implementing non-pharmaceutical interventions (NPIs) reserved for influenza pandemics (voluntary home quarantine; use of face masks by ill persons; childcare facility closures; school closures; and social distancing at schools, workplaces, and mass gatherings), and the availability and usefulness of influenza surveillance data for triggering implementation of NPIs.

**Methods:** Public health officials in all 50 states, Washington, DC, and 8 territories, and a stratified, random sample of 822 local health departments (LHDs) were surveyed in 2019.

**Results:** The response rates for the states/territories and LHDs were 75% (44/59) and 25% (206/822), respectively. About two-thirds to three-fourths of the state/territorial respondents stated that the feasibility and acceptability of implementing the NPIs were high, except for K-12 school closures lasting up to 6 weeks or 6 months. The LHD respondents also indicated that feasibility and acceptability were lowest for prolonged school closures. Compared to LHD respondents in suburban or urban areas, those in rural areas expressed lower feasibility and acceptability. Availability of influenza surveillance data in near real-time was lowest for influenza-like illness and influenza cases in schools.

**Conclusions:** The findings can provide insights regarding the implementation of NPIs during the Corona Virus Disease 2019 (COVID-19) pandemic.

## INTRODUCTION

On April 21, 2017, the Department of Health and Human Services (HHS) and the Centers for Disease Control and Prevention (CDC) released updated pre-pandemic planning guidelines entitled *Community Mitigation Guidelines to Prevent Pandemic Influenza – United States, 2017*.^1^ These guidelines replaced the 2007 interim pre-pandemic community mitigation planning guidance.^2^ The updated guidelines encourage state, tribal, local, and territorial (STLT) public health officials to plan and prepare for implementing non-pharmaceutical interventions (NPIs) early in an influenza pandemic in community settings to help slow the spread and decrease the impact of an influenza pandemic. NPIs are one of the 15 Public Health Emergency Preparedness and Response Capabilities that serve as national standards for public health preparedness planning.^3^ The 2017 guidelines delineate NPIs into two categories: 1) NPIs recommended at all times (i.e., for both seasonal influenza and influenza pandemics); and 2) NPIs reserved for influenza pandemics. Categories of NPIs recommended at all times and in all settings include personal protective measures for everyday use (voluntary home isolation of ill persons, respiratory etiquette, and hand hygiene) and environmental surface cleaning measures (routine cleaning of frequently touched surfaces and objects). During an influenza pandemic, these NPIs will be recommended regardless of the pandemic severity level. Categories of NPIs reserved for influenza pandemics include personal protective measures (voluntary home quarantine of exposed household members, and use of face masks in community settings when ill); and community measures aimed at increasing social distancing (temporarily closing or dismissing schools, limiting face-to-face contact in workplaces, and postponing or cancelling mass gatherings). During an influenza pandemic, these additional personal and community NPIs might be recommended depending on the overall pandemic severity and local conditions.

Local decisions about the selection and timing of NPIs reserved for influenza pandemics will require flexibility and modification as a pandemic progresses and new information and data become available. The 2017 guidelines include examples of surveillance data that could be used to trigger the implementation of NPIs during an influenza pandemic.^1^ In 2019, as part of ongoing pandemic influenza planning and preparedness activities, we evaluated how STLT public health officials intended to put the updated recommendations for NPIs reserved for influenza pandemics into practice in their communities. We assessed: 1) the feasibility and acceptability of and barriers to implementing the updated recommendations for NPIs reserved for influenza pandemics from the perspective of state, territorial, and local public health officials who are tasked with pre-pandemic planning, preparation, and decision-making for their respective communities; and 2) the availability and usefulness of influenza surveillance data in their jurisdictions for triggering implementation of NPIs.

## METHODS

### Study Population

The states/territories assessment comprised all 50 US states, the District of Columbia, and eight US territories and freely associated states (American Samoa, Guam, US Virgin Islands, Northern Mariana Islands, Puerto Rico, Federated States of Micronesia, Republic of the Marshall Islands, and Republic of Palau). The sampling frame for selecting LHDs comprised a universe of 2,454 LHDs – the total population of LHDs used by the National Association of County and City Health Officials (NACCHO) in their distribution of the National Profile of Local Health Departments Survey.^4^ Information on size of the population served, US Census region, and degree of urbanization of the LHDs was obtained from the NACCHO Profile data.^4^ After excluding 470 LHDs serving a population of fewer than 10,000 (which collectively serve about two percent of the total US population), 822 LHDs were sampled from 47 states; Hawaii and Rhode Island were excluded because these states did not have LHDs; and Florida was excluded as all data collection instruments distributed to LHDs in Florida must receive pre-clearance review and approval from the state health department in an effort to reduce response burden. A stratified random sample was selected from 12 strata based on the size of the population the LHD served (small = 10,000 to 49,999; medium = 50,000 to 499,999; and large = 500,000 and above) and the census region in which the LHD resided (Northeast, Midwest, South, and West). The CDC and The MayaTech Corporation determined that the project did not meet the definition of human subjects research. Data were collected under OMB Approval Number 0920-0879.

### Assessment Tool

The questionnaire covered the following four topic areas: background information on respondent and jurisdiction; status of pre-pandemic planning; feasibility and acceptability of implementing NPI recommendations during severe influenza pandemics; and availability and usefulness of influenza surveillance data for deciding when to trigger the activation of NPIs. Eight individuals from state and local health departments across the United States piloted the questionnaire in November 2018. Feedback from the pilot test resulted in minor modifications.

The topic area of feasibility and acceptability included the following eight NPIs: voluntary home quarantine; use of face masks by ill persons; temporary childcare facility closures; preemptive K- 12 school closures (for up to 2 weeks, up to 6 weeks, and up to 6 months); temporary closures of colleges and universities; social distancing measures at schools (e.g., dividing classes into smaller groups, rearranging desks so students are spaced at least 3 feet from each other); social distancing measures at workplaces (e.g., offering telecommuting, replacing in-person meetings with telephone or video conferencing, staggering work hours); and social distancing measures at mass gatherings (e.g., modifying, postponing, or canceling large events). The questions had separate four-point Likert response scales for feasibility and acceptability (high, moderately high, moderately low, low). If a respondent entered moderately low or low for feasibility or acceptability of an NPI, a text box was provided to explain the reason for their response and to describe the barriers.

The topic area of availability and usefulness of influenza surveillance data for their jurisdictions included three indicators of clinical severity of influenza (influenza-associated hospitalizations, total deaths attributed to influenza, and influenza-associated deaths among those <18 years old) and five indicators of level of influenza activity or spread (patient visits to outpatient health care providers for influenza-like illness [ILI]; proportion of respiratory specimens that test positive for influenza virus; weekly level of geographic spread of influenza; absenteeism rates due to ILI in childcare facilities, K-12 schools, or colleges and universities; and number of laboratory-confirmed influenza cases among students, teachers, and staff). The questions on the usefulness of influenza surveillance indicators had a five-point Likert response scale (extremely useful, very useful, moderately useful, slightly useful, not at all useful).

### Data Collection

Data were collected during the period from July to December 2019. An initial recruitment email was sent to public health emergency preparedness directors in the 59 state and territorial jurisdictions requesting their participation. An automated email was subsequently sent via SurveyMonkey with a link to the web-based questionnaire, with three follow-up email messages delivered 1 week apart to non-responders, resulting in 30 responses. After phone calls and up to three rounds of personalized emails were sent to non-responders, an additional 14 responses were obtained. The final response rate was 75% (44/59), with 39 states and 5 territories responding.

The *Qualtrics* survey software was used to send the web-based questionnaire to LHD preparedness coordinators and local health officials. A total of four reminder email notices were sent to non-responders, resulting in 190 responses. To increase the response rate, three additional follow-up emails were sent. Outreach efforts by NACCHO staff included an informational email to the State Associations of County and City Health Officials to inform their constituents and remind them to complete the assessment; and messages to relevant groups via e-mail, an e-newsletter, and social media. These efforts yielded approximately 16 additional responses for a final response rate of 25% (206/822).

### Analysis

The responses to the questions on feasibility were recoded to high feasibility (high feasibility + moderately high feasibility) and low feasibility (moderately low feasibility + low feasibility). Similarly, responses to the questions on acceptability were recoded to high acceptability (high acceptability + moderately high acceptability) and low acceptability (moderately low acceptability + low acceptability). A feasibility score was computed by summing the responses for the eight NPIs after assigning each NPI a score of 1 for high feasibility and a score of 0 for low feasibility. To avoid disproportionate effect of K-12 school closures/dismissals on the score, the response to closures/dismissals of up to 2 weeks was included in the score (the responses to closures/dismissals of up to 6 weeks and up to 6 months were excluded). A similar process was used to compute an acceptability score.

Because the LHDs were selected using stratified random sampling and the LHD response rate was low, sampling and non-response weights were generated using the 12 sampling strata. Among the 206 responding LHDs, 19 LHDs that provided background information but did not respond to any of the other topic areas were classified as non-responders for the purpose of computing non-response weights. PROC SURVEYFREQ, PROC SURVEYMEANS, and PROC SURVEYREG in SAS (version 9.4) were used to compute weighted percentages, weighted means, and weighted linear regression coefficients. A finite population correction factor was applied to 95% confidence intervals. For the qualitative responses on barriers (open-ended items), content analyses were conducted manually using dual-rater review.

## RESULTS

The state/territorial health department respondents comprised mainly disaster/emergency preparedness coordinators (41%), state public health officials (18%), and epidemiologists (18%). The LHD respondents were mainly local public health officials (66%) and disaster/emergency preparedness coordinators (14%). The locations of the LHDs were urban for 43%, suburban for 38%, and rural for 19%. Among the urban LHDs, the jurisdiction size was large for 15%, medium for 55%, and small for 30%; among the suburban and rural LHDs, about one-fourth were medium and three-fourths were small (none were large).

The proportion of the state/territorial respondents who reported that they were aware of or had read the updated 2017 guidelines were 93% and 82%, respectively; the corresponding proportions for the LHD respondents were 71% and 44%. Regarding incorporation of the 2017 guidelines into their pandemic influenza preparedness plans, the responses of state/territorial respondents were as follows: completed, 16%; in progress, 54%; not started, 23%; don’t know, 7%. The corresponding LHD responses were 9%, 42%, 18%, and 31%, respectively. The proportion of LHDs indicating that incorporation of the 2017 guidelines was completed or in progress was 58% for those located in urban areas, 50% for those in suburban areas, and 38% for those in rural areas.

About two-thirds to three-fourths of the state/territorial respondents stated that feasibility of implementation was high for the following NPIs: voluntary home quarantine; use of face masks by ill persons; pre-emptive closures of childcare facilities; pre-emptive closures of K-12 schools for up to 2 weeks; pre-emptive closures of colleges and universities; social distancing at schools; social distancing at workplaces; and social distancing at mass gatherings (Fig. 1). However, feasibility was perceived to be substantially lower for K-12 school closures of up to 6 weeks or 6 months (41% and 16%, respectively). For the LHDs, about 30% to 45% of respondents indicated that they did not know what the feasibility was across all NPIs (Fig. 2). However, the response pattern was similar with substantially lower feasibility for K-12 school closures of up to 6 weeks or 6 months compared to the other NPIs. The findings for acceptability were generally similar to those for feasibility (Fig. 2).

**FIGURE 1.**
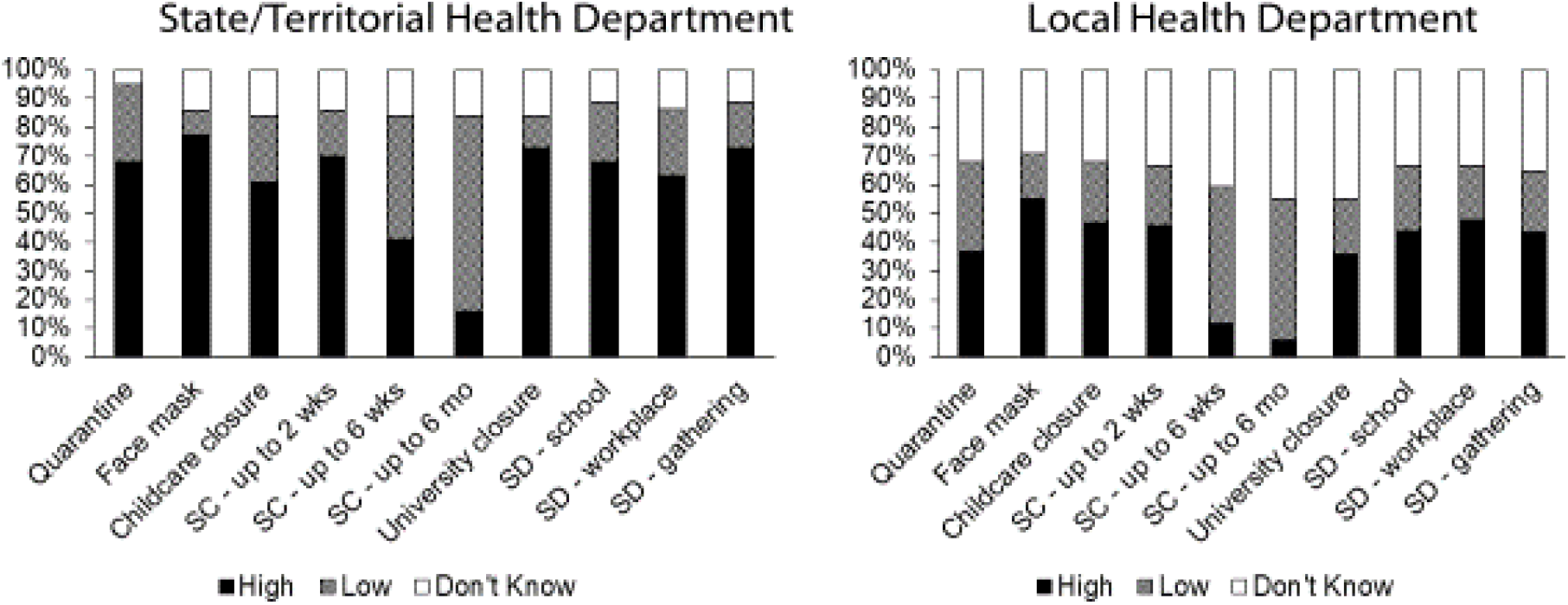
**Perceived Feasibility in State/Territorial and Local Health Department Jurisdictions of Implementing Non-Pharmaceutical Interventions during an Influenza Pandemic, 2019\*** Abbreviations: SC, school closure; SD – school, social distancing at schools; SD – workplace, social distancing at workplaces; SD – gathering, social distancing at mass gatherings (e.g., modifying, postponing, or canceling large events). *n for state/territorial and local health department jurisdictions were 44 and 187, respectively.

**FIGURE 2.**
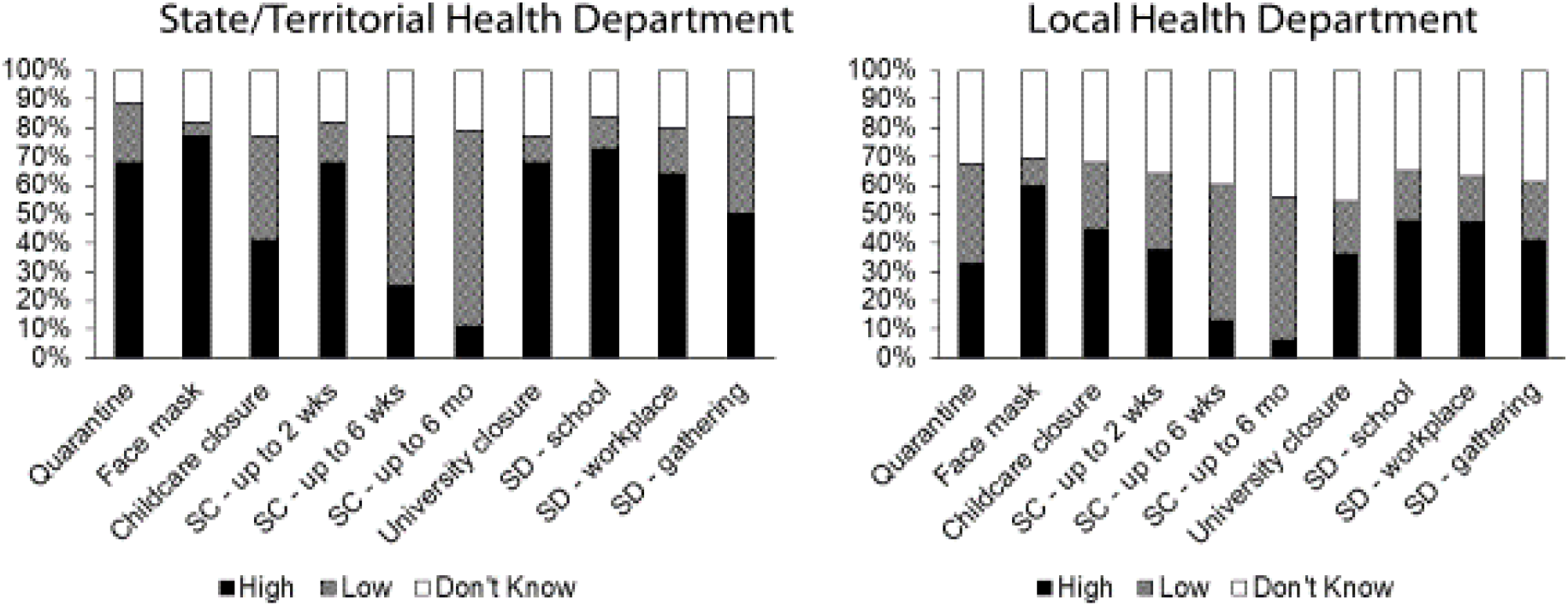
**Perceived Acceptability in State/Territorial and Local Health Department Jurisdictions of Implementing Non-Pharmaceutical Interventions during an Influenza Pandemic, 2019\*** Abbreviations: SC, school closure; SD – school, social distancing at schools; SD – workplace, social distancing at workplaces; SD – gathering, social distancing at mass gatherings (e.g., modifying, postponing, or canceling large events). *n for state/territorial and local health department jurisdictions were 44 and 187, respectively.

The feasibility and acceptability scores for the LHDs are shown in Table 1. The feasibility scores were significantly higher for urban (regression coefficient 1.02, P < 0.05) and suburban (regression coefficient 1.13, P < 0.05) LHDs compared to rural LHDs. The acceptability scores were also higher for urban and suburban LHDs than for rural LHDs.

**TABLE 1.** **Perceived Feasibility and Acceptability of Implementing Non-Pharmaceutical Interventions during an Influenza Pandemic, by Urbanicity of Local Health Department, 2019**

| Characteristics | Feasibility Score<sup>a</sup> |  |  | Acceptability Score<sup>a</sup> |  |  |
| --- | --- | --- | --- | --- | --- | --- |
|  | Unwtd n | Mean (95% CI) | Regression Coefficient (95% CI)<sup>b</sup> | Unwtd n | Mean (95% CI) | Regression Coefficient (95% CI)<sup>b</sup> |
| Overall | 144 | 4.84<br>(4.47-5.20) | - | 142 | 4.78<br>(4.40-5.16) | - |
| Urbanicity |  |  |  |  |  |  |
| Urban | 81 | 4.88<br>(4.35-5.42) | 1.02<sup>c</sup><br>(0.03-2.01) | 79 | 4.78<br>(4.27-5.29) | 1.02<br>(-0.01-2.05) |
| Suburban | 41 | 5.15<br>(4.58-5.72) | 1.13<sup>c</sup><br>(0.09-2.17) | 41 | 5.12<br>(4.41-5.83) | 1.14<sup>c</sup><br>(0.01-2.27) |
| Rural | 20 | 4.14<br>(3.26-5.03) | 0<br>(Referent) | 20 | 4.15<br>(3.25-5.05) | 0<br>(Referent) |

Abbreviations: Unwtd, unweighted; CI, confidence interval.
<sup>a</sup>Feasibility and acceptability scores, each ranging from 0 to 8, were computed by summing the responses to eight questions on feasibility and the corresponding eight questions on acceptability (excluding the questions on school closures for up to 6 weeks and school closures for up to 6 months) (high = 1; low = 0; do not know/not sure/blank = missing). Jurisdictions with missing responses on all eight questions (43 for feasibility and 45 for acceptability) were excluded. Information on urbanicity was missing for two jurisdictions.
<sup>b</sup>Linear regression models were run separately for feasibility score and acceptability score (dependent variables). The independent variables in the models were urbanicity and census region. Jurisdiction size was dropped from the models because of collinearity with urbanicity.
<sup>c</sup>P < 0.05.

The barriers to implementing NPIs are listed in Tables A1 to A12 of the Appendix. Among state/territorial and LHD respondents that rated the feasibility and acceptability of implementing NPI recommendations as moderately low or low, the financial impact of the recommendations on individuals, businesses, and the community was a recurring theme of barriers reported. Barriers to prolonged school closures (up to 6 weeks, up to 6 months) indicated that the financial burden was particularly tied to employment issues (e.g., inability to miss work and limited childcare options, inability to telework); other barriers included loss of school meals for vulnerable children and disruption of education. Barriers for rural areas included difficulty in enforcement (e.g., quarantine, childcare facility closures), people not wanting government interfering with their lives (e.g., quarantine), and families not having adequate resources for distance learning (K- 12 school closures or dismissals). Ratings of low acceptability implicated an anticipated lack of buy-in by both the public and community leaders, and challenges with achieving compliance.

Figure 3 shows the availability of influenza surveillance data that might provide information for triggering implementation of NPIs. For the states/territories, about half of the jurisdictions reported having near real-time data on outpatient ILI visits, geographic spread of influenza cases, proportion of specimens positive for influenza, influenza-associated hospitalizations, and influenza deaths in children; about one-third reported having near real-time data on total influenza-associated deaths; and about 10% reported having near real-time data on ILI-related absenteeism and influenza cases in schools. For the LHDs, about 30% to 40% reported that they did not know whether near real-time data were available for the surveillance indicators for their jurisdiction. For the state/territorial and LHD respondents who had near real-time data, most of the respondents indicated that the indicators were extremely useful or very useful for deciding when to trigger the activation of NPIs in their jurisdictions (Tables 2 and 3).

**TABLE 2.**
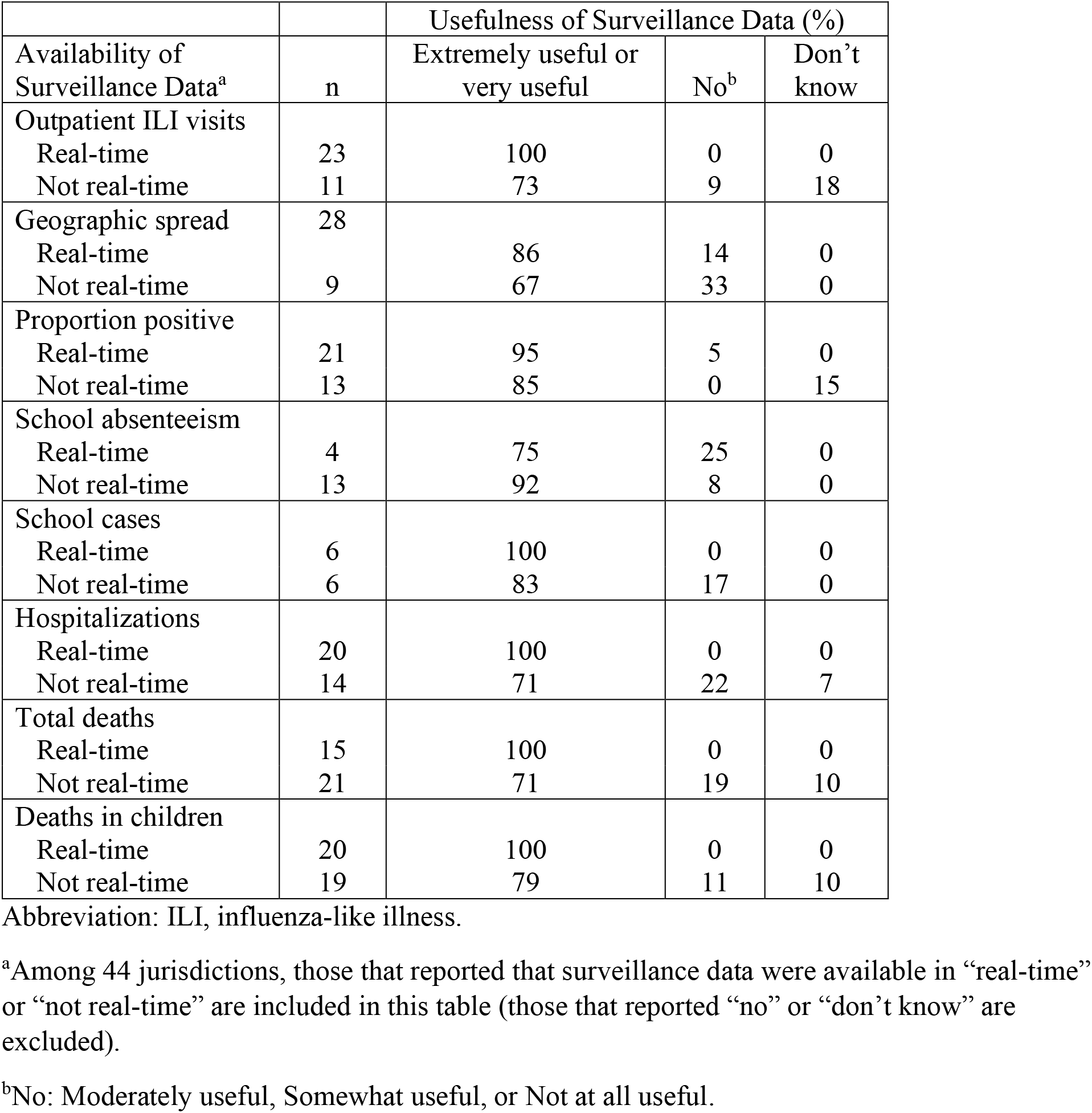
**State/Territorial Health Department Perceptions of Usefulness of Surveillance Data for Deciding When to Trigger Implementation of Non-Pharmaceutical Interventions during an Influenza Pandemic, by Timeliness of Data, 2019**

**TABLE 3.** **Local Health Department Perceptions of Usefulness of Surveillance Data for Deciding When to Trigger Implementation of Non-Pharmaceutical Interventions during an Influenza Pandemic, by Timeliness of Data, 2019**

|  |  | Usefulness of Surveillance Data<br>(weighted %) |  |  |
| --- | --- | --- | --- | --- |
| Availability of<br>Surveillance Data<sup>a</sup> | Unweighted<br>n | Extremely useful<br>or very useful | No<sup>b</sup> | Don't<br>know |
| Outpatient ILI visits |  |  |  |  |
| Real-time | 40 | 74 | 24 | 2 |
| Not real-time | 40 | 63 | 27 | 10 |
| Geographic spread |  |  |  |  |
| Real-time | 51 | 82 | 18 | 0 |
| Not real-time | 44 | 78 | 18 | 4 |
| Proportion positive |  |  |  |  |
| Real-time | 46 | 96 | 4 | 0 |
| Not real-time | 50 | 77 | 19 | 4 |
| School absenteeism |  |  |  |  |
| Real-time | 36 | 91 | 9 | 0 |
| Not real-time | 49 | 64 | 27 | 9 |
| School cases |  |  |  |  |
| Real-time | 23 | 100 | 0 | 0 |
| Not real-time | 34 | 68 | 30 | 2 |
| Hospitalizations |  |  |  |  |
| Real-time | 61 | 92 | 8 | 0 |
| Not real-time | 50 | 70 | 17 | 13 |
| Total deaths |  |  |  |  |
| Real-time | 47 | 98 | 2 | 0 |
| Not real-time | 62 | 68 | 22 | 10 |
| Deaths in children |  |  |  |  |
| Real-time | 53 | 96 | 3 | 1 |
| Not real-time | 61 | 71 | 25 | 4 |

Abbreviation: ILI, influenza-like illness.
<sup>a</sup>Among 187 jurisdictions, those that reported that surveillance data were available in “real-time” or “not real-time” are included in this table (those that reported “no” or “don’t know” are excluded).
<sup>b</sup>No: Moderately useful, Somewhat useful, or Not at all useful.

**FIGURE 3.**
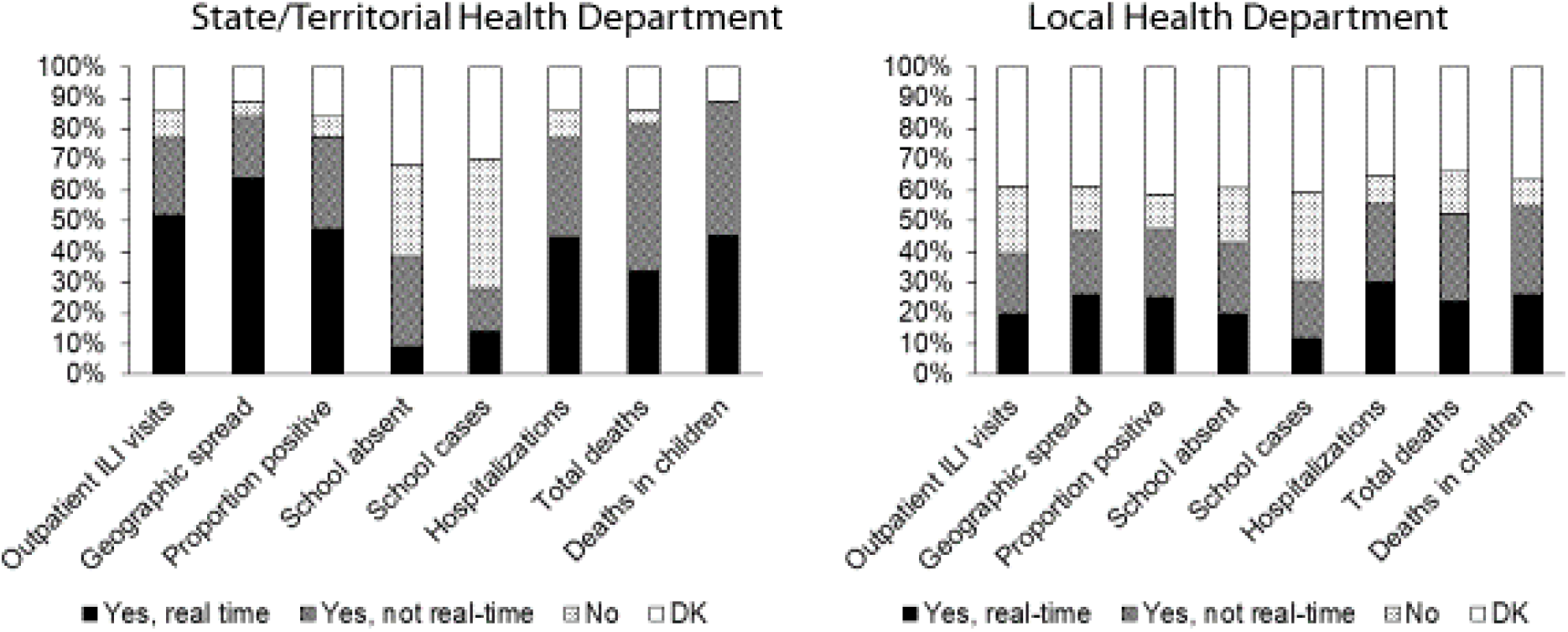
**Availability of Surveillance Data in State/Territorial and Local Health Department Jurisdictions for Triggering Implementation of Non-Pharmaceutical Interventions during an Influenza Pandemic, 2019\*** Abbreviations: ILI, influenza-like illness; DK, don’t know. *n for state/territorial and local health department jurisdictions were 44 and 187, respectively.

## DISCUSSION

About two-thirds to three-fourths of the state/territorial respondents stated that the feasibility and acceptability of implementing the NPIs reserved for influenza pandemics were high, except for prolonged K-12 school closures. The LHD respondents also indicated that feasibility and acceptability were lowest for prolonged school closures. The feasibility and acceptability scores were lower for LHDs located in rural areas than those in suburban or urban areas. Availability of influenza surveillance data in near real-time was lowest for ILI absenteeism rates and influenza cases in schools.

Our findings on perceived NPI acceptability, feasibility, and barriers are consistent with those of previous studies. A study conducted in 2006 indicated that most individuals would comply with community mitigation recommendations during a severe influenza pandemic.^5^ A national survey of adults during the 2009 H1N1 influenza pandemic showed high public approval for government recommendations related to school closures (80%), avoiding places where many people gather (69%), and wearing masks in public (71%).^6^ A survey of public health officials in 50 US states and eight territories and freely associated states in 2015 indicated that 85 percent of the jurisdictions had or did not need the legal authority to temporarily close child care facilities, K–12 schools, or colleges/universities, or cancel mass gatherings.^7^ About two-thirds of state/territorial respondents in our evaluation indicated that feasibility of social distancing in K- 12 schools was high. A previous report indicated that within-school social distancing practices were generally more feasible for elementary schools than secondary schools; for reduced-schedule practices, shortening the school week for the entire school was more feasible than shortening the school day.^8^ Our evaluation found that feasibility and acceptability were lowest for prolonged K-12 school closures, and that barriers included inability to work, loss of income, missing school meals, and continuity of education. A previous study has reported that the social and economic effects of school closures include loss of income for parents who may have to stay home to take care of their children, difficulties sustaining teaching and learning, and loss of school meals for underprivileged children who rely on free or reduced price school lunches.^9^ Another study reported that a substantial proportion of adults would face severe financial problems if they had to stay home from work for several weeks to comply with community mitigation recommendations, with a disproportionate effect for persons with lower incomes and for racial and ethnic minorities.^5^ A study found that working adults would be less able to comply if they were unable to work from home or did not have paid sick leave.^10^

We found that feasibility and acceptability scores were lower for LHDs located in rural areas. This finding is consistent with a previous report that that social distancing orders were issued less often in rural areas in response to communicable disease outbreaks.^11^ Evaluations conducted in 2020 during the coronavirus disease 2019 (COVID-19) pandemic reported higher use of cloth face coverings in urban compared to rural areas^12^ and lower adoption of stay-at-home orders in states with higher proportion of rural residents.^13^

We found that availability of influenza surveillance data was lowest for ILI absenteeism rates and influenza cases in schools. This may be because these two indicators are not a part of the US Influenza Surveillance System.^14^ School absenteeism data collected by school districts are not standardized and rarely include information about the illness that caused the absence.^15^ Lack of data on ILI absenteeism and influenza cases in schools may hamper the ability to decide when to trigger proactive school closures.^16^ A survey of LHDs in 2015 indicated that the most common concern about the use of social distancing (including quarantine, isolation, school closures, and work closures) was the magnitude of public health impact; other concerns included legal, political, financial, and sociocultural issues, and the impacts to vulnerable populations.^11^ A survey administered in 2015 to 62 Public Health Emergency Preparedness directors in the 50 US states, eight US territories and freely associated states, and four cities indicated that the most important factors for selecting and triggering the implementation of NPIs during an influenza pandemic were severity of illness, transmissibility, and populations most affected.^7^ Other important factors were CDC and subject matter expert recommendations, geographic spread of the disease, disease impact in relation to available mitigation resources, and vaccine availability.

Our evaluation has some limitations. First, although we requested that respondents consult with colleagues if necessary, the responses may not be reflective of the perspective of the entire health department. Second, the LHD response rate was low, as in another recent evaluation of LHD preparedness.^11^ However, our use of non-response weight in order to align the responding sample to the original sample in terms of jurisdiction size and census region might have reduced bias. Finally, because we did not have the names of the jurisdictions in the state/territorial and LHD analytic datasets to preserve respondents’ confidentiality, we could not conduct an in-depth assessment of geographic variability.

Our data collection was completed just 1 month before the first cases of Corona Virus Disease 2019 (COVID-19) were reported in China and the disease subsequently spread around the world. Because of the severity of the COVID-19 pandemic, NPIs that were implemented during the spring of 2020 in the United States included stay-at-home orders, business closures, and K-12 school closures for several months.^13, 17, 18^ Most K-12 public schools that closed offered distance learning and meal services for students^19^ and about 45 percent of the general population worked from home instead of their normal workplaces.^20^ The US government provided economic assistance to American workers and businesses, and required covered employers to provide paid sick leave or expanded family and medical leave if an employee was unable to work because of COVID-19 illness or quarantine or to take care of a quarantined family member or a child whose school or child care provider was closed.^21, 22^

## CONCLUSIONS

To our knowledge, our assessment is the first national-scope investigation to systematically evaluate perceived NPI feasibility, acceptability, and barriers and the availability and usefulness of influenza surveillance data for triggering implementation of NPIs by surveying all state/territorial health departments and a nationally representative sample of LHDs. The results of our assessment were intended to help inform NPI implementation considerations 2 years after release of the updated 2017 guidelines. The findings can provide insights regarding the implementation of NPIs during the COVID-19 pandemic.

## Data Availability

Data can be shared upon request.

## ACKNOWLEDGEMENTS

We would like to acknowledge the contributions of the project’s stakeholder engagement group consisting of representatives from the Association of State and Territorial Health Officials (ASTHO), Council of State and Territorial Epidemiologists (CSTE), National Association of County and City Health Officials (NACCHO), and the National Public Health Information Coalition (NPHIC); MayaTech’s internal evaluation support team – Jamie Weinstein, Barbara Draley, and Mitch Wang – and their subject matter expert, Barbara Goldrick; and fellow CDC project team members – Neha Kanade, Jasmine Kenney, and Tiffani Phelps. We also would like to acknowledge that the data collection for the local health department assessment was implemented by NACCHO under the direction of Dr. Debra Dekker and her team.

## DISCLAIMER

The findings and conclusions in this report are those of the authors and do not necessarily represent the official position of the Centers for Disease Control and Prevention.

## APPENDIX

**TABLE A1.** Voluntary Home Quarantine during an Influenza Pandemic: Reasons/Barriers for Rating the Feasibility of this Recommendation as Moderately Low or Low, 2019.

| Thematic Category | States and Territories (N=13) | Local Health Departments (N=37) |
| --- | --- | --- |
| Disruptive to ability to work (may not get paid depending on job). The number of people without paid sick leave or unable to work from home | ✓ | ✓ |
| Economics: Most families need two incomes to survive, reluctant to miss work because of lost wages | ✓ | ✓ |
| Rural composition of state/sparse population density would make compliance and enforcement problematic | ✓ | ✓ |
| People (especially those in rural communities) do not want “government” interfering with their lives | ✓ | ✓ |
| People will be reluctant to miss work if they are NOT sick | ✓ | ✓ |
| There will be a significant portion of the public who will not follow the recommendation | ✓ |  |
| Health literacy: Not fully understanding voluntary home quarantine |  | ✓ |
| Communal spaces and population density in urban areas (homeless shelters, jails) |  | ✓ |

N represents number of respondents (unweighted).

**TABLE A2.** Voluntary Home Quarantine during an Influenza Pandemic: Reasons/Barriers for Rating the Acceptability of this Recommendation as Moderately Low or Low, 2019.

| Thematic Categories | State and Territories (N=9) | Local Health Departments (N=53) |
| --- | --- | --- |
| People do not like or are not used to having their movements restricted | ✓ | ✓ |
| Negative impact on low-income/low-wage parents | ✓ | ✓ |
| Many will feel the need to go to work, especially those living paycheck to paycheck. Barriers are financial. | ✓ | ✓ |
| Lack of education and understanding regarding transmissibility | ✓ |  |
| Unavailability of childcare during the school year | ✓ |  |
| Variances between jurisdictions accepting closures | ✓ |  |
| Enforcement would be difficult due to geography (rural) | ✓ |  |
| Depends on how people perceive the severity of the pandemic and likelihood of becoming ill | ✓ |  |
| People (especially those in rural communities) do not want “government” interfering with their lives |  | ✓ |
| Businesses and services having strict leave policies and that keeps workers from being able to home quarantine |  | ✓ |
| People do not want to miss work, especially if they are not ill |  | ✓ |

**TABLE A3.** Use of Face Masks by Ill Persons during an Influenza Pandemic: Reasons/Barriers for Rating the Feasibility of this Recommendation as Moderately Low or Low, 2019.

| Thematic Category | States and Territories (N=3) | Local Health Departments (N=29) |
| --- | --- | --- |
| Identifying ill persons and providing them with face masks | ✓ |  |
| Supply of masks (potential shortage of masks) | ✓ |  |
| Prioritizing masks for healthcare workers if there is a shortage | ✓ |  |
| Face mask distribution (will people purchase their own, are they being distributed to certain populations?) | ✓ |  |
| Public mistrust of the government |  | ✓ |
| Masks being perceived as uncomfortable |  | ✓ |
| Belief that masks not only used for the ill, but those who are well (not ill) |  | ✓ |
| Consistency in training on how to use the masks, this may be difficult to implement & enforce |  | ✓ |

**TABLE A4.**
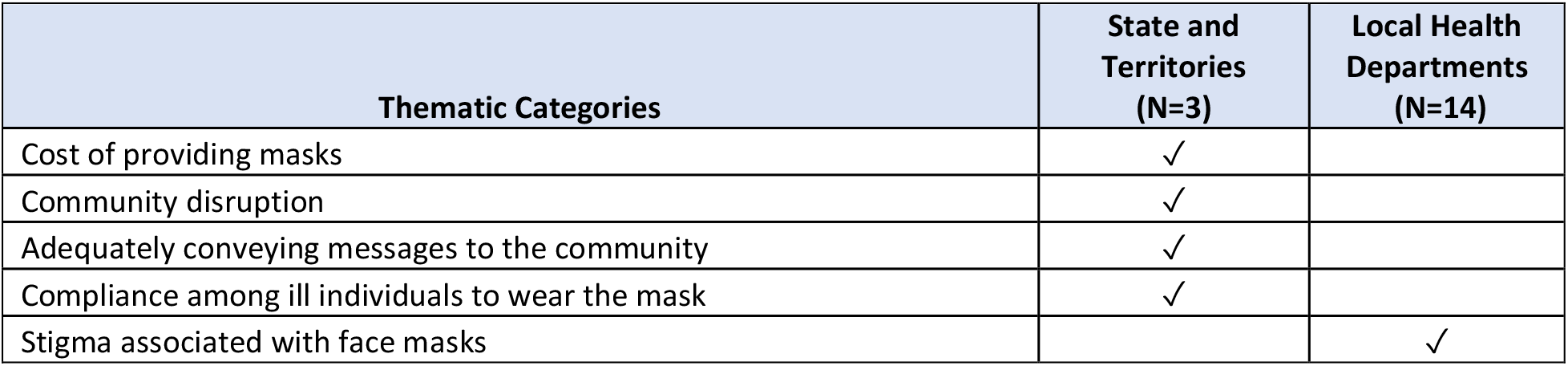
Use of Face Masks by Ill Persons during an Influenza Pandemic: Reasons/Barriers for Rating the Acceptability of this Recommendation as Moderately Low or Low, 2019.

| Thematic Categories | State and Territories (N=3) | Local Health Departments (N=14) |
| --- | --- | --- |
| Cost of providing masks | ✓ |  |
| Community disruption | ✓ |  |
| Adequately conveying messages to the community | ✓ |  |
| Compliance among ill individuals to wear the mask | ✓ |  |
| Stigma associated with face masks |  | ✓ |

**TABLE A5.**
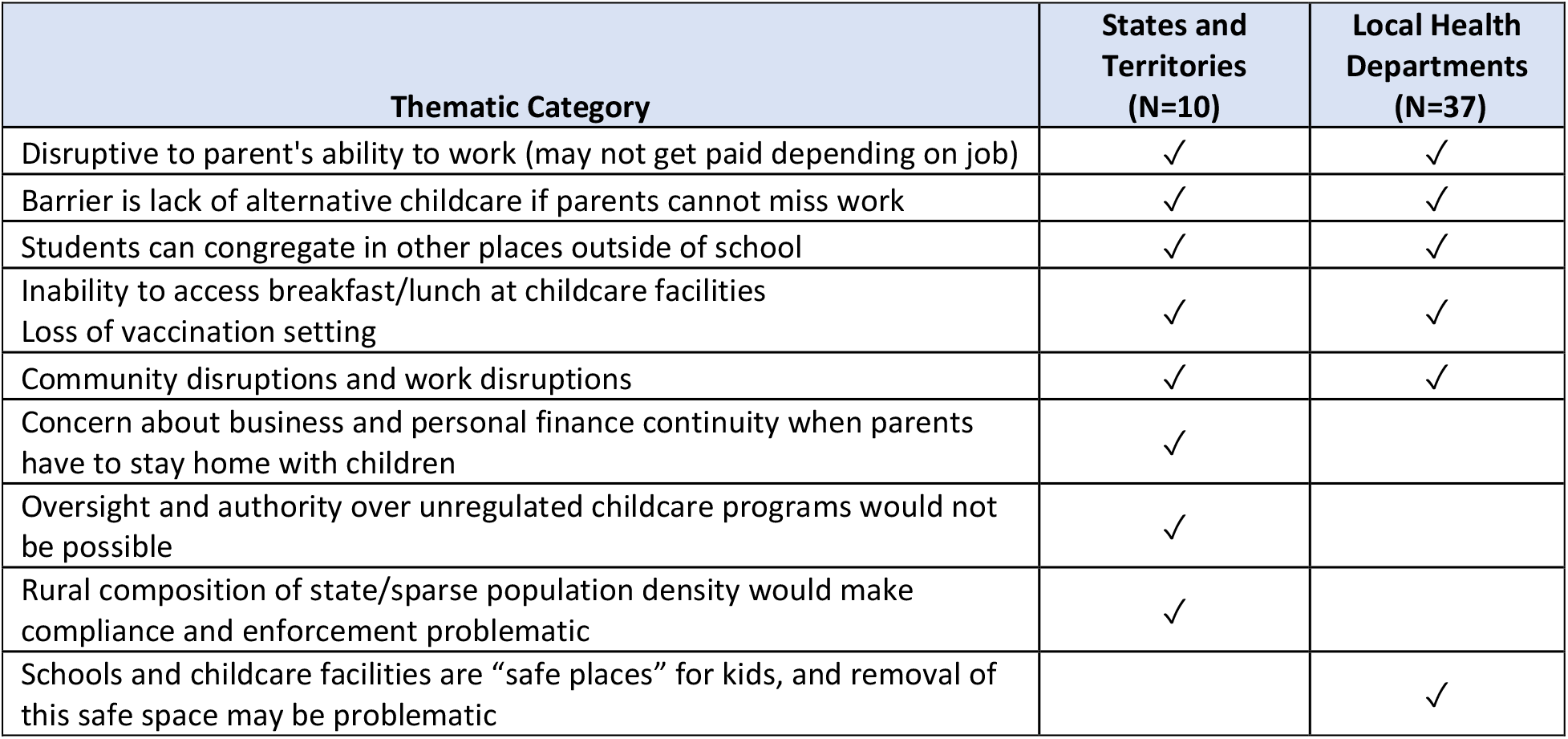
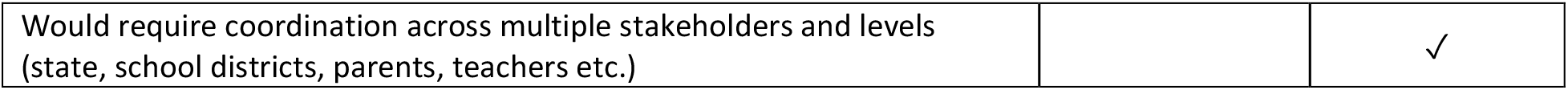
Childcare Facility Closures or Dismissals during an Influenza Pandemic: Reasons/Barriers for Rating the Feasibility of this Recommendation as Moderately Low or Low, 2019.

**TABLE A6.** Childcare Facility Closures or Dismissals during an Influenza Pandemic: Reasons/Barriers for Rating the Acceptability of this Recommendation as Moderately Low or Low, 2019.

| Thematic Categories | State and Territories (N=16) | Local Health Departments (N=42) |
| --- | --- | --- |
| Loss of income generated for childcare facilities | ✓ | ✓ |
| Lack of alternative childcare for working parents | ✓ | ✓ |
| Negative impact on daycare staff | ✓ | ✓ |
| Negative impact on the general community | ✓ | ✓ |
| State and local school districts and boards do not always follow public health recommendations | ✓ | ✓ |
| Depends on how people perceive the severity of the pandemic and likelihood of becoming ill | ✓ | ✓ |
| Variances between jurisdictions accepting closures | ✓ | ✓ |
| Parents need to work | ✓ |  |
| Would be difficult to enforce with unlicensed programs | ✓ |  |
| Negative impact on partner workforce | ✓ |  |
| Negative impact on low-income/low-wage parents | ✓ |  |
| Lost ability to access breakfast/lunch | ✓ |  |
| Hard to use disease surveillance to time the recommendation for closure | ✓ |  |
| If these facilities close, ad hoc operators will emerge thereby negating the intended results. | ✓ |  |
| Limiting freedoms (high rate of unimmunized and people who do not like being told what to do) |  | ✓ |

**TABLE A7.** Preemptive K-12 School Closures or Dismissals during an Influenza Pandemic: Reasons/Barriers for Rating the Feasibility of this Recommendation as Moderately Low or Low, 2019.

| Thematic Category<br>*[timeframe of closures, if included]* | States and Territories (N=30) | Local Health Departments (N=37) |
| --- | --- | --- |
| Barrier is lack of alternative childcare if parents cannot miss work<br>*[2 weeks, up to 6 weeks, 6 months]* | ✓ | ✓ |
| Disruptive to parents' ability to work (may not get paid depending on job)<br>*[2 weeks, up to 6 weeks, 6 months]* | ✓ |  |
| Community disruptions and work disruptions (impact on local economy, loss in tax revenues) *[6 months]* | ✓ | ✓ |
| Children to rely on free/reduced-cost breakfast/lunch at school would lose access *[up to 6 weeks, 6 months]* | ✓ | ✓ |
| Learning lags in children's education. Not all parents/school districts are equipped to move to online/web-based teaching and learning *[up to 6 weeks, 6 months]* | ✓ | ✓ |
| Concern about business and personal finance continuity when parents have to stay home with children | ✓ |  |
| Special needs of low-income parents and their children |  | ✓ |

| Thematic Category<br>*[timeframe of closures, if included]* | States and Territories<br>(N=30) | Local Health Departments<br>(N=37) |
| --- | --- | --- |
| Low-income families and some rural families may not have adequate resources for distance learning |  | ✓ |

**TABLE A8.** **Preemptive K-12 School Closures or Dismissals during an Influenza Pandemic: Reasons/Barriers for Rating the Acceptability of this Recommendation as Moderately Low or Low, 2019**

| Thematic Categories | State and Territories<br>(N=30) | Local Health Departments<br>(N=81) |
| --- | --- | --- |
| Perceived level of threat or the odds of contracting the illness | ✓ | ✓ |
| Lack of alternative childcare | ✓ | ✓ |
| Financial impact/economic sustainability (personal and business) | ✓ | ✓ |
| Parents' inability to take significant time off work | ✓ |  |
| The longer the timeframe for the closure, the less accepted it will be |  | ✓ |

**TABLE A9.**
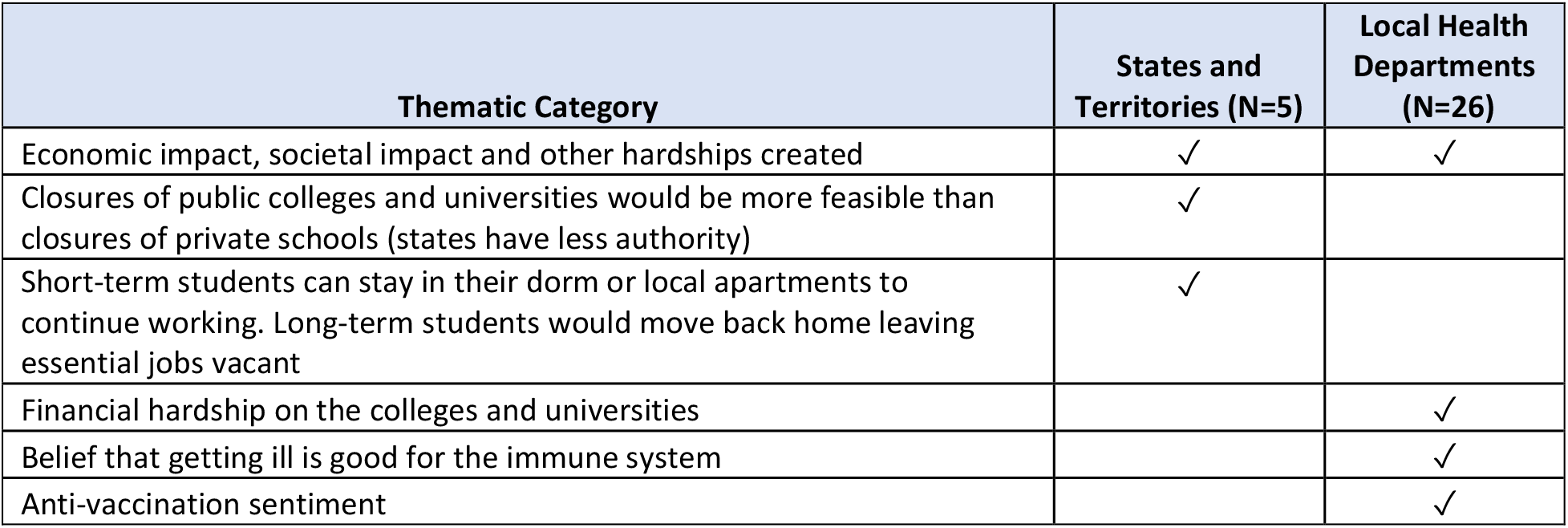
**Temporary Closures or Dismissals of Colleges and Universities during an Influenza Pandemic: Reasons/Barriers for Rating the Feasibility of this Recommendation as Moderately Low or Low, 2019**

| Thematic Category | States and Territories (N=5) | Local Health Departments<br>(N=26) |
| --- | --- | --- |
| Economic impact, societal impact and other hardships created | ✓ | ✓ |
| Closures of public colleges and universities would be more feasible than closures of private schools (states have less authority) | ✓ |  |
| Short-term students can stay in their dorm or local apartments to continue working. Long-term students would move back home leaving essential jobs vacant | ✓ |  |
| Financial hardship on the colleges and universities |  | ✓ |
| Belief that getting ill is good for the immune system |  | ✓ |
| Anti-vaccination sentiment |  | ✓ |

**TABLE A10.**
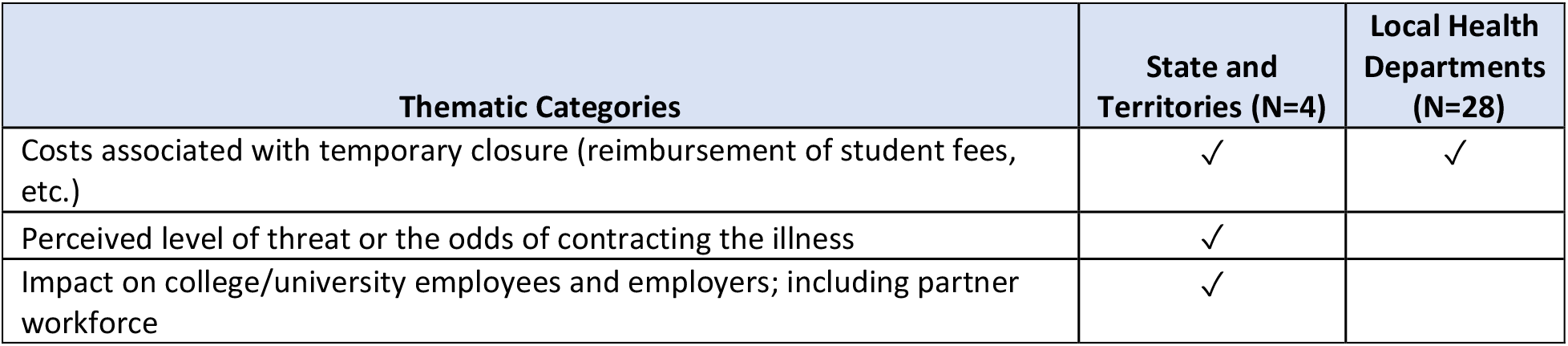
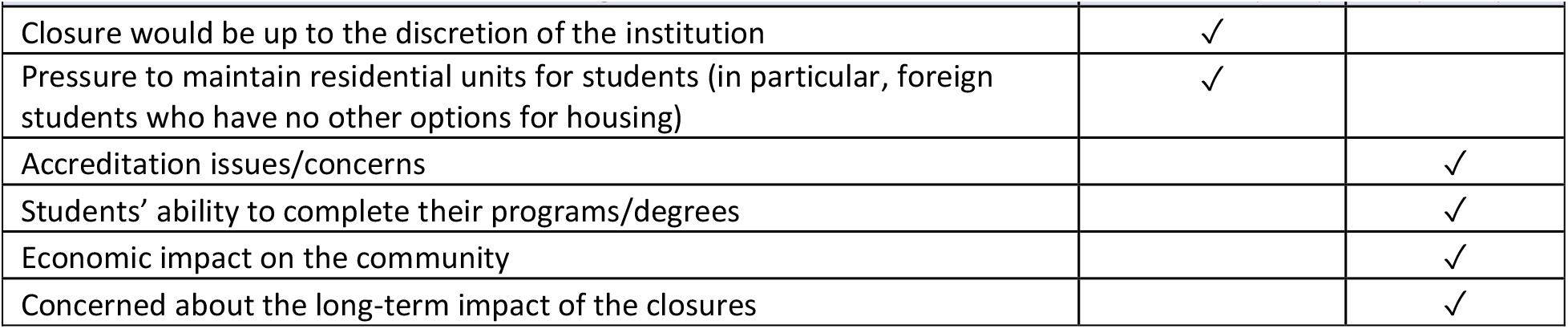
**Temporary Closures or Dismissals of Colleges and Universities during an Influenza Pandemic: Reasons/Barriers for Rating the Acceptability of this Recommendation as Moderately Low or Low, 2019**

**TABLE A11.**
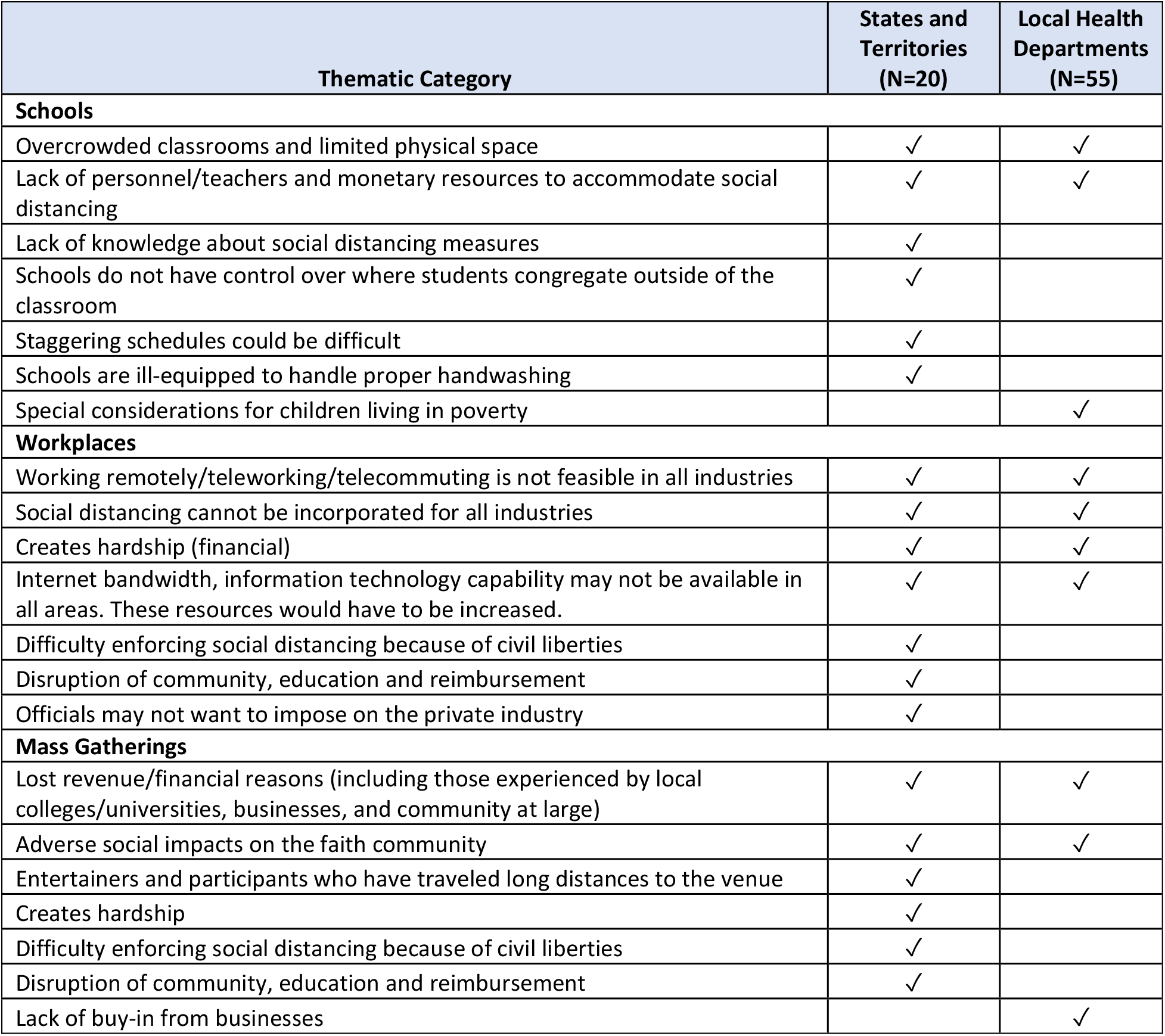
Social Distancing during an Influenza Pandemic: Reasons/Barriers for Rating the Feasibility of this Recommendation as Moderately Low or Low, 2019.

| Thematic Category | States and Territories (N=20) | Local Health Departments (N=55) |
| --- | --- | --- |
| **Schools** |  |  |
| Overcrowded classrooms and limited physical space | ✓ | ✓ |
| Lack of personnel/teachers and monetary resources to accommodate social distancing | ✓ | ✓ |
| Lack of knowledge about social distancing measures | ✓ |  |
| Schools do not have control over where students congregate outside of the classroom | ✓ |  |
| Staggering schedules could be difficult | ✓ |  |
| Schools are ill-equipped to handle proper handwashing | ✓ |  |
| Special considerations for children living in poverty |  | ✓ |
| **Workplaces** |  |  |
| Working remotely/teleworking/telecommuting is not feasible in all industries | ✓ | ✓ |
| Social distancing cannot be incorporated for all industries | ✓ | ✓ |
| Creates hardship (financial) | ✓ | ✓ |
| Internet bandwidth, information technology capability may not be available in all areas. These resources would have to be increased. | ✓ | ✓ |
| Difficulty enforcing social distancing because of civil liberties | ✓ |  |
| Disruption of community, education and reimbursement | ✓ |  |
| Officials may not want to impose on the private industry | ✓ |  |
| **Mass Gatherings** |  |  |
| Lost revenue/financial reasons (including those experienced by local colleges/universities, businesses, and community at large) | ✓ | ✓ |
| Adverse social impacts on the faith community | ✓ | ✓ |
| Entertainers and participants who have traveled long distances to the venue | ✓ |  |
| Creates hardship | ✓ |  |
| Difficulty enforcing social distancing because of civil liberties | ✓ |  |
| Disruption of community, education and reimbursement | ✓ |  |
| Lack of buy-in from businesses |  | ✓ |

**TABLE A12.** Social Distancing: Reasons/Barriers for Rating the Acceptability of this Recommendation as Moderately Low or Low.

| Thematic Category | States and Territories (N=20) | Local Health Departments (N=47) |
| --- | --- | --- |
| **Schools** |  |  |
| Staggering school schedules which is less effective than closures, and the push back associated with closing schools | ✓ |  |
| Economic impact on schools that cannot fill classrooms |  | ✓ |
| Adverse impact on parents' paychecks |  | ✓ |
| Inadequate capacity to monitor and enforce the recommendation |  | ✓ |
| Pushback from community members when sporting events/large school events are cancelled | ✓ |  |
| **Workplaces** |  |  |
| Lack of adequate messaging to persuade businesses |  | ✓ |
| Economic impact on employees |  | ✓ |
| Inability to enforce recommendation |  | ✓ |
| **Mass Gatherings** |  |  |
| Cancelling sporting events can have political ramifications | ✓ |  |
| Public resistance to cancelled sporting events could have political concerns | ✓ |  |
| Faith leaders not buying in |  | ✓ |
| Lack of effective messaging to get the public to comply with the recommendation |  | ✓ |
| Financial implications for employees in mass gathering settings |  | ✓ |

